# Proportional Mortality and Years of Potential Life Lost Due to Liver Diseases among Agricultural Workers, Brazil, 2017 to 2022

**DOI:** 10.1101/2024.07.09.24309605

**Authors:** Jailma dos Santos Silva, Soraia Arruda, Thayane Silva Nunes, Wiler de Paula Dias, Adedayo Michael Awoniyi, Armando Meyer, Cleber Cremonese

**Affiliations:** Institute of Collective Health, Federal University of Bahia, Salvador, Brazil; Porto Alegre Clinical Hospital. Epidemiological Surveillance, Porto Alegre, Brazil; Environmental and Occupational Health Branch, Public Health Institute, Federal University of Rio de Janeiro, Rio de Janeiro, Brazil

**Keywords:** Liver Diseases, Occupational Mortality, Years of Potential Life Lost, Rural Workers, Health Information System

## Abstract

The objective of the study was to describe the profile and calculate the Years of Potential Life Lost (YPLL) due to liver diseases in Brazilian agricultural workers, between 2017 and 2022. For this, we analyzed microdata available in the Mortality Information System (Sistema de Informação de Mortalidade - SIM), considering the outcome as the underlying cause of death with codes K70-K77 (ICD-10). Workers’ profile was characterized according to socio-demographic variables and the Brazilian regions, with a comparison group formed by all other Brazilian workers aged 18 to 69 who died in the same period and from the same underlying cause. Calculations of proportional mortality, YPLL rates, and YPLL rate ratios were applied. In the studied period, 15,362 deaths due to liver diseases were recorded among Brazilian agricultural workers, with an average age at death of 51.3 years (±10.7), concentrated in K70 - Alcoholic liver disease (53.8%). Higher proportional mortality was observed in men (86.2%), Brown race (61.1%), up to 49 years old (40.9%), with ≤7 years of education level (52.4%), and residents in Northeast (56.9%). The total sum of YPLL was 382,869 years among agricultural workers, with YPLL rate of 4,527 years per 100,000 workers and YPLL rate ratio 1.45 times higher than the national average. The concentration of deaths due to K70 raises concern for potential chronic exposure to alcoholic beverages. These results highlight the early causes of deaths resulting from liver diseases among agricultural workers, especially those in the Northeast region of Brazil and of Brown race.

## INTRODUCTION

Liver diseases encompass a range of morbidities that impair the liver’s ability to effectively perform its functions such as substance and nutrient metabolism, blood filtration, toxin elimination, bile production, and protein synthesis^1^. The International Classification of Diseases (ICD-10)^2^ categorizes liver diseases under eight codes (K70-K77), which can result from viral infections, intoxication by harmful substances, excessive alcohol consumption, poor diet, autoimmune conditions, and genetic factors^3–6^.

Annually, liver diseases are responsible for two million deaths globally, making it the eleventh leading cause of death and accounting for 4% of the total mortality. Two-thirds of these deaths occur in men^7^. Factors such as low education level and non-white race/ethnicity (hereafter referred to as race) background have been linked to higher incidences of liver diseases^8–10^.

In Low- and Middle-Income Countries (LMICs) like Brazil, factors such as sex, race, and geographic location, combined with risk behaviors and limited access to healthcare, exacerbate liver dysfunction, increasing morbidity and mortality rates^4,6,11^. In Brazil, from 1996 to 2012, the mortality rate from liver diseases showed an upward trend, reaching 20 deaths per 100,000 people, although with somewhat fluctuations averaging 19 deaths per 100,000 Brazilians over the past decade. During this period, all regions exhibited a rising trend in these deaths, with the Northeast and Midwest regions particularly notable for a higher proportion of K70-Alcoholic liver disease. Liver diseases account for 3% of total deaths in Brazil, costing the Brazilian healthcare services approximately USD 60 million annually^12^.

Although previous studies have provided valuable insights into the epidemiological assessment of global morbidity and mortality ^5,6,11^ as well as national liver diseases^12^, investigations often overlook specific segments such as economic activity or type of occupation. Agricultural workers represent a distinct labor group that frequently handles pesticides and heavy metals, endures long working hours, and performs intense physical labor. Numerous studies have examined the adverse health effects on agricultural workers, focusing on chronic diseases such as neoplasms^13–16^, cardiovascular diseases^17–19^, degenerative diseases^20–22^, and common mental disorders^15,23,24^. Additionally, acute effects, including work accidents^25^ and exogenous intoxications^16,26,27^ are well-documented. However, studies evaluating the relationship between agricultural occupations and liver diseases, considering factors like demographics, geographic location, occupational exposure, and access to preventive examinations or treatments are rare or nonexistent in Brazil.

As a result, this study aims to characterize the profile of death records from liver diseases and calculate the Years of Potential Life Lost (YPLL) among Brazilian agricultural workers compared to other workers, aged 18 to 69 years, taking into account sociodemographic determinants and geographical distribution across Brazil from 2017 to 2022.

## METHODS

### Study design and population

This is a descriptive study that analyzed microdata from death records due to liver diseases, in individuals aged 18 to 69 years across the entire Brazilian territory, from 2017 to 2022. The final dataset includes all death records that provide information on both the cause of death and the occupation of the deceased.

### Data collection and processing

The data were obtained from the Mortality Information System (Sistema de Informação sobre Mortalidade - SIM), derived from Death Certificates (DCs) provided by the Department of Informatics of the Unified Health System (Departamento de Informática do Sistema Único de Saúde - DATASUS), accessible through https://datasus.saude.gov.br. For outcome selection, all records from field 40 of the DC, (Causa básica – CAUSABAS), containing codes K70 to K77 referring to Liver Diseases according to Chapter XI - Diseases of the Digestive System, as per ICD-10^2^ were considered. Briefly, these codes include K70-Alcoholic liver disease, K71-Toxic liver disease, K72-Unspecified liver failure, K73-Chronic hepatitis, K74-Fibrosis and cirrhosis of the liver, K75-Other inflammatory liver diseases, K76-Other diseases of the liver, and K77-Liver disorders in diseases classified elsewhere.

The usual occupation indicated in field 14 (Ocupação – OCUP), refers to the type of work the deceased mostly engaged in during their productive life, as per the Death Certificate Manual^28^. The Brazilian Classification of Occupations (Classificação Brasileira de Ocupações - CBO)^29^ was used to categorize ‘Agricultural workers’, encompassing codes 61 (Agricultural producers), 62 (Agricultural workers), and 64 (Agricultural and forestry mechanization workers). All other valid occupation codes were grouped under ‘other workers’.

The workers’ population data were obtained from the Demographic Census available in the IBGE Automatic Recovery System (SIDRA), specifically table 3584 selecting the age range of 18 to 69 years (https://sidra.ibge.gov.br/Tabela/3584)^30^.

To characterize the profile of deaths resulting from liver diseases, we analyzed information on sex (male/female), age groups (18 - 29, 30 - 39, 40 - 49, 50 - 59, and 60 - 69 years), race (White, Black, Yellow, Brown, and Indigenous), years of education (≤3, 4 - 7, 8 - 11, and ≥12 years), marital status (single, married, legally separated, and widowed), regions (North, Northeast, Midwest, Southeast, and South), and municipality typology (urban, adjacent intermediary, remote intermediary, adjacent rural, and remote rural). Missing data are presented in the descriptive tables. All analyses were performed in R software version 4.1.2^31^. All datasets and codes used during this study are available in Zenodo under Creative Commons 4.0 license, accessible through https://doi.org/10.5281/zenodo.1209302332.

### Data analysis

The studied populations were characterized using absolute values and their corresponding frequencies. Proportional Mortality (PM) was calculated by dividing the total number of deaths in the category of interest by the total number of deaths in the variable of interest for the period multiplied by 100.

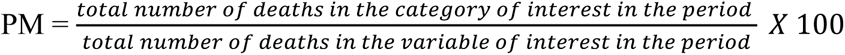

The mean age of death (MAoD) was calculated by dividing the sum of ages at death by the total number of deaths in the variable of interest.

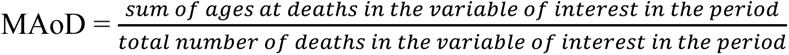

To calculate the YPLL, we considered the difference between the mean age of the variable of interest and the corresponding Life Expectancy (LE - 76.2 years in 2019, overall for Brazil) (mi)^33^, multiplied by the total number of deaths in the variable of interest (ai).

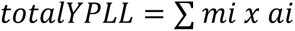

The percentage distribution of YPLL (%YPLL) by categories of interest was calculated by dividing the YPLL in the category of interest by the total YPLL in the variable of interest, multiplied by 100.

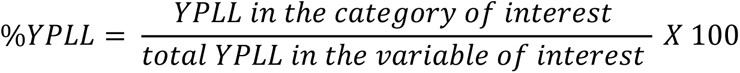

The YPLL rate (YPLLr) was calculated by dividing the YPLL by the standard population, multiplied by 100,000 workers. Here, YPLL rates were calculated by Brazilian region, by agricultural workers, and other workers.

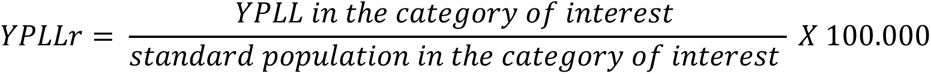

Finally, the YPLLr ratios were calculated for the regions and groups of workers. For regions analysis, those with the lowest YPLL rates were used as the reference (denominator) and “other workers” used as reference (denominator) for occupation groups.

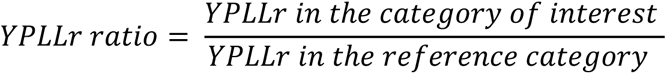

Inferential analyses were not applied due to the census nature of the data, which includes all deaths registered in the investigation period, thereby eliminating the need for hypothesis testing.

### Ethical Statement

Given the study used non-identifiable open access data (microdata) available in the public domain, ethical approval was not required for this study.

## RESULTS

There were a total of 4,058,039 deaths registered in SIM among adults aged 18 to 69 years during the study period. Among these, 120,552 cases (2.9%) were attributed to liver diseases as the underlying cause of death (ICD-10, K70-K77). In general, the MAoD was 52.2 years (±13.7), whereas for liver diseases, it was 53.5 years (±10.3). Deaths due to liver diseases were significantly higher (*p-value<0.001*) among men (80.4%), in the age group of 50 to 59 years (34.5%), of Brown race (46.3%), and single individuals (40.1%). Geographically, the Southeast and Northeast regions accounted for 43.7% and 28.9% of these deaths, and urban municipalities 76.1% respectively (Table 1).

**Table 1.**
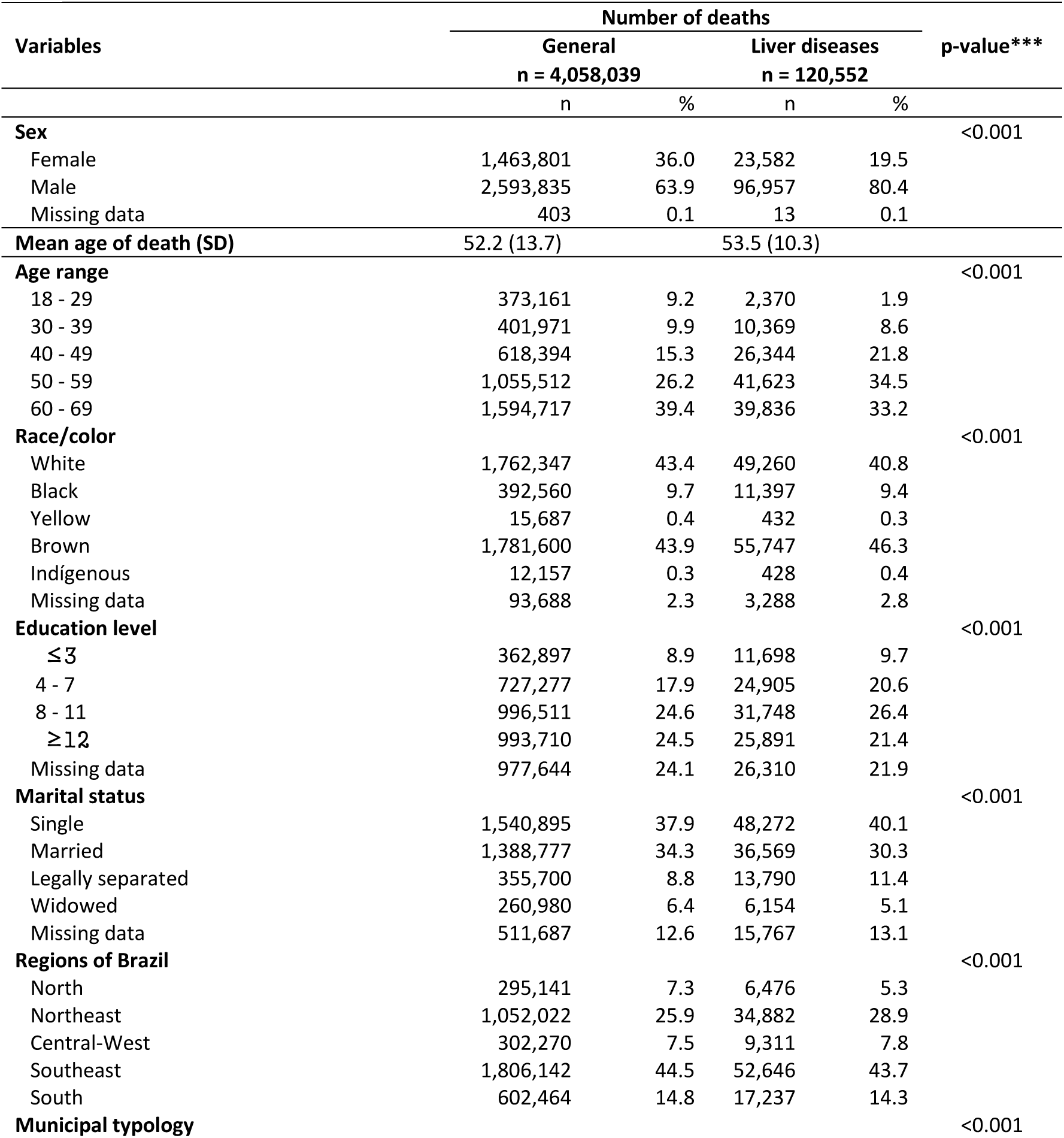
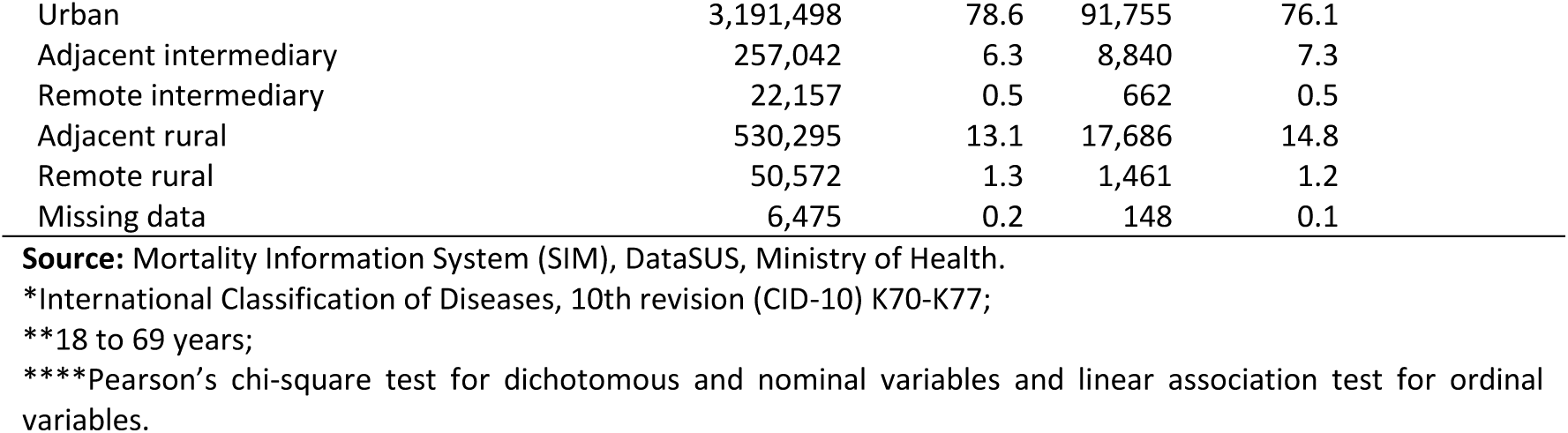
Distribution of general deaths and those resulting from liver diseases* among adult Brazilian populations** from 2017 to 2022.

| Variables | Number of deaths |  |  |  | p-value*** |
| --- | --- | --- | --- | --- | --- |
|  | General |  | Liver diseases |  |  |
|  | n = 4,058,039 |  | n = 120,552 |  |  |
|  | n | % | n | % |  |
| Sex |  |  |  |  | <0.001 |
| Female | 1,463,801 | 36.0 | 23,582 | 19.5 |  |
| Male | 2,593,835 | 63.9 | 96,957 | 80.4 |  |
| Missing data | 403 | 0.1 | 13 | 0.1 |  |
| Mean age of death (SD) | 52.2 (13.7) |  | 53.5 (10.3) |  |  |
| Age range |  |  |  |  | <0.001 |
| 18 - 29 | 373,161 | 9.2 | 2,370 | 1.9 |  |
| 30 - 39 | 401,971 | 9.9 | 10,369 | 8.6 |  |
| 40 - 49 | 618,394 | 15.3 | 26,344 | 21.8 |  |
| 50 - 59 | 1,055,512 | 26.2 | 41,623 | 34.5 |  |
| 60 - 69 | 1,594,717 | 39.4 | 39,836 | 33.2 |  |
| Race/color |  |  |  |  | <0.001 |
| White | 1,762,347 | 43.4 | 49,260 | 40.8 |  |
| Black | 392,560 | 9.7 | 11,397 | 9.4 |  |
| Yellow | 15,687 | 0.4 | 432 | 0.3 |  |
| Brown | 1,781,600 | 43.9 | 55,747 | 46.3 |  |
| Indigenous | 12,157 | 0.3 | 428 | 0.4 |  |
| Missing data | 93,688 | 2.3 | 3,288 | 2.8 |  |
| Education level |  |  |  |  | <0.001 |
| ≤3 | 362,897 | 8.9 | 11,698 | 9.7 |  |
| 4 - 7 | 727,277 | 17.9 | 24,905 | 20.6 |  |
| 8 - 11 | 996,511 | 24.6 | 31,748 | 26.4 |  |
| ≥12 | 993,710 | 24.5 | 25,891 | 21.4 |  |
| Missing data | 977,644 | 24.1 | 26,310 | 21.9 |  |
| Marital status |  |  |  |  | <0.001 |
| Single | 1,540,895 | 37.9 | 48,272 | 40.1 |  |
| Married | 1,388,777 | 34.3 | 36,569 | 30.3 |  |
| Legally separated | 355,700 | 8.8 | 13,790 | 11.4 |  |
| Widowed | 260,980 | 6.4 | 6,154 | 5.1 |  |
| Missing data | 511,687 | 12.6 | 15,767 | 13.1 |  |
| Regions of Brazil |  |  |  |  | <0.001 |
| North | 295,141 | 7.3 | 6,476 | 5.3 |  |
| Northeast | 1,052,022 | 25.9 | 34,882 | 28.9 |  |
| Central-West | 302,270 | 7.5 | 9,311 | 7.8 |  |
| Southeast | 1,806,142 | 44.5 | 52,646 | 43.7 |  |
| South | 602,464 | 14.8 | 17,237 | 14.3 |  |
| Municipal typology |  |  |  |  | <0.001 |

|  |  |  |  |  |
| --- | --- | --- | --- | --- |
| Urban | 3,191,498 | 78.6 | 91,755 | 76.1 |
| Adjacent intermediary | 257,042 | 6.3 | 8,840 | 7.3 |
| Remote intermediary | 22,157 | 0.5 | 662 | 0.5 |
| Adjacent rural | 530,295 | 13.1 | 17,686 | 14.8 |
| Remote rural | 50,572 | 1.3 | 1,461 | 1.2 |
| Missing data | 6,475 | 0.2 | 148 | 0.1 |

**Source:** Mortality Information System (SIM), DataSUS, Ministry of Health.
\*International Classification of Diseases, 10th revision (CID-10) K70-K77;
\*\*18 to 69 years;
\*\*\*Pearson's chi-square test for dichotomous and nominal variables and linear association test for ordinal variables.

Agricultural workers reported younger MAoD (about 3 years younger) compared to other workers (Table 2). Regarding PM, male agricultural workers represented 86.2% of cases, as against 78.2% of other workers, while Brown race individuals accounted for 61.1% of deaths among agricultural workers, as against 43.1% among other workers. Notably, the Northeast region of the country reported 56.9% of liver disease-related deaths among agricultural workers, which was 2.7 times higher than cases registered among other workers in the same region (Table 2).

**Table 2.**
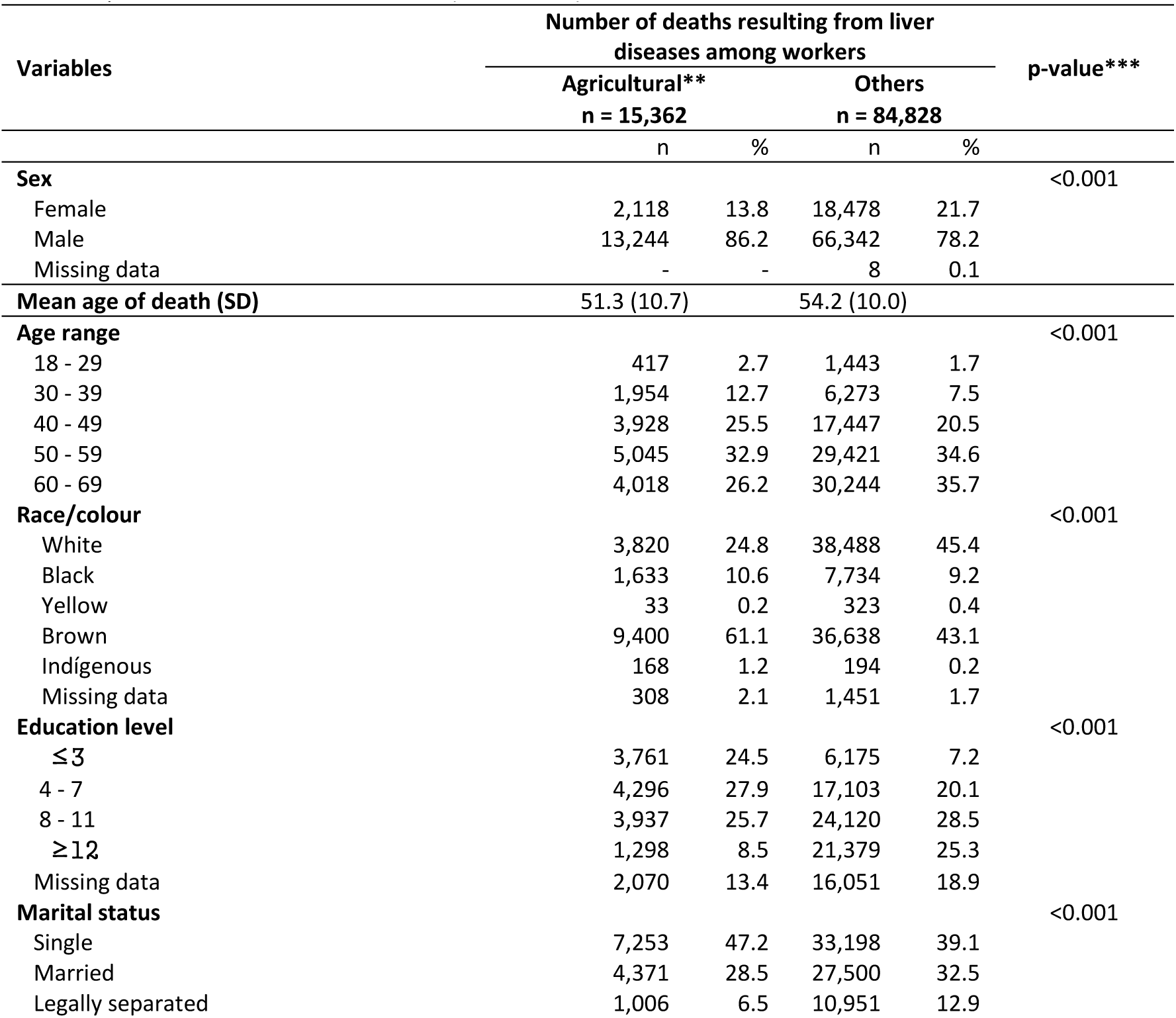
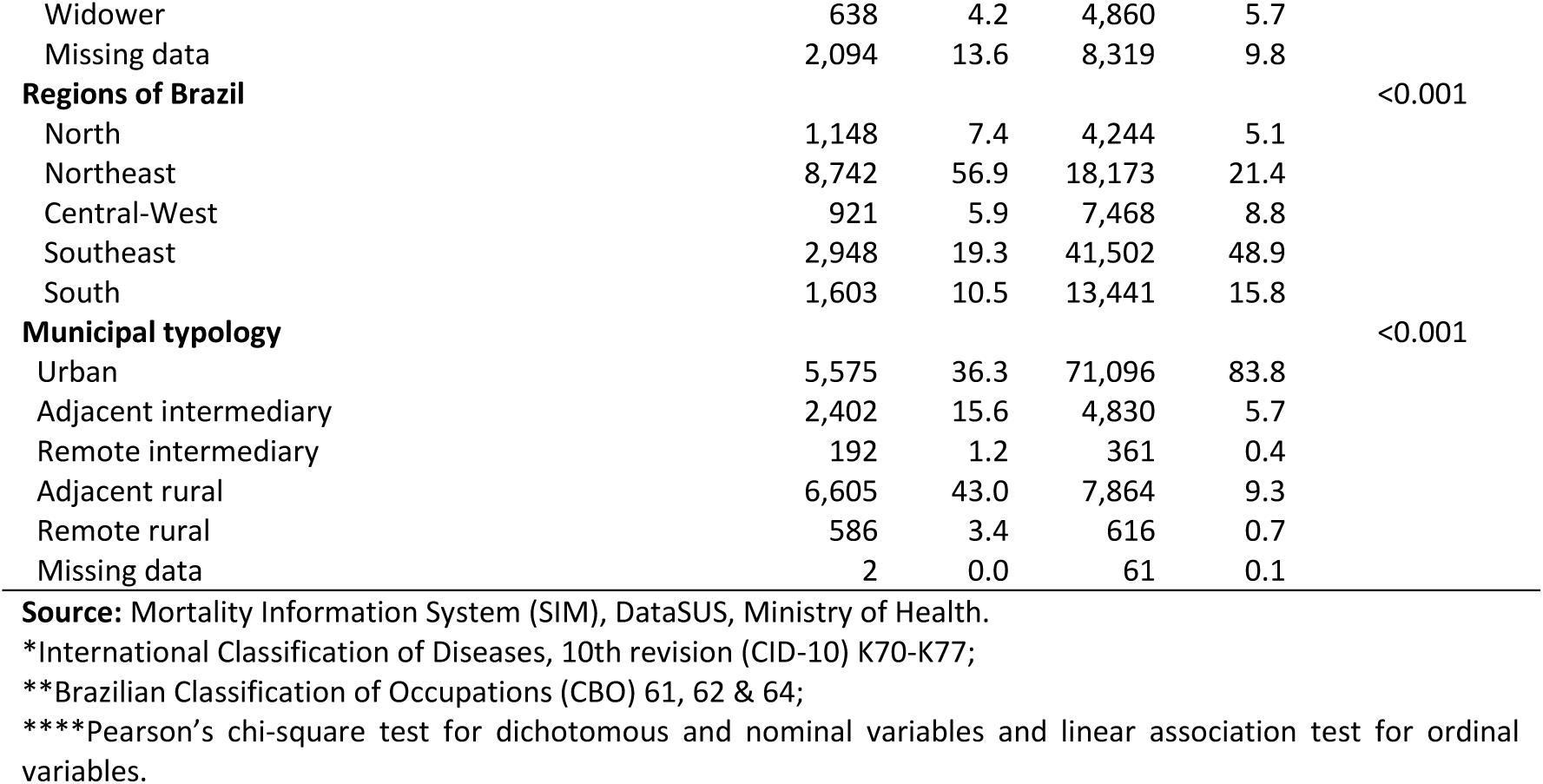
Distribution of deaths resulting from liver diseases* among agricultural and non-agricultural workers aged 18 to 69 years in Brazil from 2017 to 2022. (n = 100,190)

| Variables | Number of deaths resulting from liver diseases among workers |  |  |  | p-value*** |
| --- | --- | --- | --- | --- | --- |
|  | Agricultural** |  | Others |  |  |
|  | n = 15,362 |  | n = 84,828 |  |  |
|  | n | % | n | % |  |
| **Sex** |  |  |  |  | <0.001 |
| Female | 2,118 | 13.8 | 18,478 | 21.7 |  |
| Male | 13,244 | 86.2 | 66,342 | 78.2 |  |
| Missing data | - | - | 8 | 0.1 |  |
| **Mean age of death (SD)** | 51.3 (10.7) |  | 54.2 (10.0) |  |  |
| **Age range** |  |  |  |  | <0.001 |
| 18 - 29 | 417 | 2.7 | 1,443 | 1.7 |  |
| 30 - 39 | 1,954 | 12.7 | 6,273 | 7.5 |  |
| 40 - 49 | 3,928 | 25.5 | 17,447 | 20.5 |  |
| 50 - 59 | 5,045 | 32.9 | 29,421 | 34.6 |  |
| 60 - 69 | 4,018 | 26.2 | 30,244 | 35.7 |  |
| **Race/colour** |  |  |  |  | <0.001 |
| White | 3,820 | 24.8 | 38,488 | 45.4 |  |
| Black | 1,633 | 10.6 | 7,734 | 9.2 |  |
| Yellow | 33 | 0.2 | 323 | 0.4 |  |
| Brown | 9,400 | 61.1 | 36,638 | 43.1 |  |
| Indigenous | 168 | 1.2 | 194 | 0.2 |  |
| Missing data | 308 | 2.1 | 1,451 | 1.7 |  |
| **Education level** |  |  |  |  | <0.001 |
| ≤3 | 3,761 | 24.5 | 6,175 | 7.2 |  |
| 4 - 7 | 4,296 | 27.9 | 17,103 | 20.1 |  |
| 8 - 11 | 3,937 | 25.7 | 24,120 | 28.5 |  |
| ≥12 | 1,298 | 8.5 | 21,379 | 25.3 |  |
| Missing data | 2,070 | 13.4 | 16,051 | 18.9 |  |
| **Marital status** |  |  |  |  | <0.001 |
| Single | 7,253 | 47.2 | 33,198 | 39.1 |  |
| Married | 4,371 | 28.5 | 27,500 | 32.5 |  |
| Legally separated | 1,006 | 6.5 | 10,951 | 12.9 |  |
| Widower | 638 | 4.2 | 4,860 | 5.7 |  |
| Missing data | 2,094 | 13.6 | 8,319 | 9.8 |  |
| **Regions of Brazil** |  |  |  |  | <0.001 |
| North | 1,148 | 7.4 | 4,244 | 5.1 |  |
| Northeast | 8,742 | 56.9 | 18,173 | 21.4 |  |
| Central-West | 921 | 5.9 | 7,468 | 8.8 |  |
| Southeast | 2,948 | 19.3 | 41,502 | 48.9 |  |
| South | 1,603 | 10.5 | 13,441 | 15.8 |  |
| **Municipal typology** |  |  |  |  | <0.001 |
| Urban | 5,575 | 36.3 | 71,096 | 83.8 |  |
| Adjacent intermediary | 2,402 | 15.6 | 4,830 | 5.7 |  |
| Remote intermediary | 192 | 1.2 | 361 | 0.4 |  |
| Adjacent rural | 6,605 | 43.0 | 7,864 | 9.3 |  |
| Remote rural | 586 | 3.4 | 616 | 0.7 |  |
| Missing data | 2 | 0.0 | 61 | 0.1 |  |

**Source:** Mortality Information System (SIM), DataSUS, Ministry of Health.
\*International Classification of Diseases, 10th revision (CID-10) K70-K77;
\*\*Brazilian Classification of Occupations (CBO) 61, 62 & 64;
\*\*\*\*Pearson's chi-square test for dichotomous and nominal variables and linear association test for ordinal variables.

Among the 8 subdivisions of liver diseases in the ICD-10, alcoholic liver disease (K70) had the highest PM in both groups of workers. In agricultural workers, it accounted for 53.8% of deaths in this subcategory, while in other workers, it accounted for 44.9%. The K74 subgroup, fibrosis and cirrhosis of the liver, showed the second highest PM, with 28.1% among agricultural workers and 34.3% among other workers (Table 3).

**Table 3.** Distribution of deaths resulting from liver diseases, K70-K77 subgroups of chapter XI of CID-10 among agricultural and non-agricultural workers aged 18 to 69 years in Brazil from 2017 to 2022. (n = 100,190)

| CID-10, Chapter XI – Diseases of the digestive system, K70-K77 subgroup | Number of deaths among workers |  |  |  |
| --- | --- | --- | --- | --- |
|  | Agricultural* |  | Others |  |
|  | n = 15,362 |  | n = 84,828 |  |
|  | n | % | n | % |
| K70 – Alcoholic liver disease | 8,276 | 53.8 | 38,122 | 44.9 |
| K71 – Toxic liver disease | 93 | 0.6 | 554 | 0.6 |
| K72 – Liver failure not classified elsewhere | 1,152 | 7.5 | 6,760 | 7.9 |
| K73 – Chronic hepatitis not classified elsewhere | 96 | 0.6 | 376 | 0.5 |
| K74 – Liver fibrosis and cirrhosis | 4,308 | 28.1 | 28,992 | 34.3 |
| K75 – Other inflammatory liver diseases | 206 | 1.3 | 1,842 | 2.2 |
| K76 – Other liver diseases | 1,231 | 8.1 | 8,182 | 9.5 |
| K77 – Liver disorders in diseases classified elsewhere | - | - | - | - |

**Source:** Mortality Information System (SIM), DataSUS, Ministry of Health.
\*Brazilian Classification of Occupations (CBO) 61, 62 & 64; CID-10 = International Classification of Diseases, 10th revision

In general, agricultural workers had lower MAoDs, except for K75 and the Central-West region. Alcoholic liver disease (K70) recorded the lowest MAoD among agricultural workers with 50.1 years (±10.4), compared to 52.8 years (±9.7) among other workers. Among agricultural workers, lower MAoDs were also observed among males with 51.1 years (±10.6), individuals of Black race 50.3 years (±10.8), Brown race 50.7 years (±10.7), Indigenous 49.0 years (±12.1), single individuals 47.7 years (±10.6), and in Northeast region 50.1 years (±10.9) (Table 4).

**Table 4.** Years of Potential Life Lost (YPLL) due to liver diseases, K70-K77 subgroups of chapter XI of CID-10, among agricultural and non-agricultural workers* aged 18 to 69 years in Brazil from 2017 to 2022. (n = 15,362 agricultural; n = 84,828 others)

| Variables | Mean age of death (SD) |  | YPLL |  | % YPLL |  |
| --- | --- | --- | --- | --- | --- | --- |
|  | Agricultural | Others | Agricultural | Others | Agricultural | Others |
| **K70-K77\*\* Subgroups** |  |  |  |  |  |  |
| K70 | 50.1 (10.4) | 52.8 (9.7) | 216,003.6 | 737,380.8 | 56.4 | 47.2 |
| K71 | 50.7 (13.2) | 52.6 (13.2) | 2,371.5 | 10,761.6 | 0.6 | 0.6 |
| K72 | 51.2 (11.7) | 54.0 (11.4) | 28,800 | 124,764 | 7.5 | 7.9 |
| K73 | 51.3 (12.4) | 56.0 (10.0) | 2,390.4 | 6,100.4 | 0.6 | 0.4 |
| K74 | 52.7 (10.4) | 56.0 (9.3) | 101,238 | 495,526.2 | 26.4 | 31.8 |
| K75 | 53.2 (11.8) | 52.1 (13.6) | 4,738 | 38,560 | 1.3 | 2.5 |
| K76 | 54.0 (11.1) | 55.0 (10.5) | 27,328.2 | 149,990 | 7.2 | 9.6 |
| K77 | - | - | - | - | - | - |
| **Sex** |  |  |  |  |  |  |
| Female | 52.9 (11.1) | 54.7 (11.0) | 49,349.4 | 397,277 | 13.3 | 21.4 |
| Male | 51.1 (10.6) | 54.1 (9.8) | 322,424.4 | 1,466,158.2 | 86.7 | 78.7 |
| Missing data | - | 53.5 (12.5) | - | 181.6 | - | 0.9 |
| **Age range** |  |  |  |  |  |  |
| 18 - 29 | 25.8 (2.7) | 25.1 (3.2) | 21,016.8 | 73,737.3 | 5.5 | 3.9 |
| 30 - 39 | 35.4 (2.7) | 35.6 (2.7) | 79,723.2 | 254,683.8 | 20.8 | 13.7 |
| 40 - 49 | 44.8 (2.8) | 45.0 (2.8) | 123,339.2 | 544,346.4 | 32.6 | 29.2 |
| 50 - 59 | 54.4 (2.8) | 54.7 (2.8) | 109,981 | 632,551.5 | 28.6 | 33.9 |
| 60 - 69 | 64.2 (2.8) | 64.3 (2.8) | 48,216 | 359,903.6 | 12.5 | 19.3 |
| **Race/colour** |  |  |  |  |  |  |
| White | 53.3 (10.2) | 55.6 (9.5) | 87,478 | 785,155.2 | 22.8 | 42.3 |
| Black | 50.3 (10.8) | 53.4 (10.3) | 42,294.7 | 176,335.2 | 11.1 | 9.5 |
| Yellow | 52.2 (10.5) | 55.5 (10.6) | 792 | 6,686.1 | 0.3 | 0.4 |
| Brown | 50.7 (10.7) | 53.0 (10.3) | 239,700 | 850,001.6 | 62.7 | 45.7 |
| Indigenous | 49.0 (12.1) | 50.0 (11.9) | 4,569.6 | 5,082.8 | 1.2 | 0.3 |
| Missing data | 51.8 (10.7) | 54.4 (10.0) | 7,515.2 | 31,631.8 | 1.9 | 1.8 |
| **Education level** |  |  |  |  |  |  |
| ≤3 | 54.0 (9.8) | 55.8 (9.8) | 83,494.2 | 125,970 | 21.8 | 6.7 |
| 4 - 7 | 51.5 (10.5) | 55.2 (9.5) | 106,111.2 | 359,163 | 27.7 | 19.2 |
| 8 - 11 | 50.0 (10.8) | 53.7 (10.0) | 103,149.4 | 542,700 | 26.9 | 29.2 |
| ≥12 | 47.8 (11.5) | 53.1 (10.4) | 36,863.2 | 493,854.9 | 9.7 | 26.5 |
| Missing data | 50.7 (10.7) | 54.9 (10.0) | 52,785 | 341,886.3 | 13.9 | 18.4 |
| **Marital status** |  |  |  |  |  |  |
| Single | 47.7 (10.6) | 49.7 (10.5) | 206,710.5 | 879,747 | 53.9 | 47.1 |
| Married | 55.6 (9.1) | 57.7 (8.3) | 90,042.6 | 508,750 | 23.6 | 27.3 |
| Legally separated | 54.8 (8.6) | 56.7 (7.8) | 21,528.4 | 213,544.5 | 5.7 | 11.5 |
| Widower | 59.2 (8.0) | 61.2 (6.6) | 10,846 | 72,900 | 2.9 | 3.9 |
| Missing data | 50.6 (10.5) | 53.2 (10.0) | 53,606.4 | 191,337 | 13.9 | 10.2 |
| **Regions of Brazil** |  |  |  |  |  |  |
| North | 52.5 (11.0) | 53.2 (10.9) | 27,207.6 | 97,612 | 7.2 | 5.3 |
| Northeast | 50.1 (10.9) | 53.0 (10.6) | 228,166.2 | 421,613.6 | 59.6 | 22.6 |
| Central-West | 53.5 (9.8) | 52.6 (10.2) | 20,906.7 | 176,244.8 | 5.4 | 9.4 |
| Southeast | 52.1 (10.1) | 54.9 (9.7) | 71,046.8 | 883,992.6 | 18.7 | 47.4 |
| South | 54.3 (9.5) | 54.9 (9.5) | 35,105.7 | 286,293.3 | 9.1 | 15.3 |
| Brazil | 51.3 (10.7) | 54.2 (10.0) | 382,513.8 | 1,866,216 | - | - |
| **Municipal typology** |  |  |  |  |  |  |
| Urban | 52.6 (10.4) | 54.5 (9.9) | 131,570 | 1,542,783.2 | 34.7 | 82.9 |
| Adjacent intermediary | 51.1 (10.5) | 53.1 (10.4) | 60,290.2 | 111,573 | 15.9 | 5.9 |
| Remote intermediary | 48.8 (11.5) | 51.2 (11.9) | 5,260.8 | 9,025 | 1.4 | 0.4 |
| Adjacent rural | 51.1 (10.5) | 53.0 (10.8) | 165,785.5 | 182,444.8 | 43.7 | 9.8 |
| Remote rural | 49.7 (11.9) | 51.5 (11.3) | 15,529 | 15,215.2 | 4.1 | 0.9 |
| Missing data | 59.5 (0.7) | 51.2 (10.3) | 33.4 | 1,525 | 0.2 | 0.1 |

**Source:** Mortality Information System (SIM), DataSUS, Ministry of Health.
\* Brazilian Classification of Occupations (CBO) 61, 62 & 64;
\*\*K70-Alcoholic liver disease; K71-Toxic liver disease; K72-Liver failure not classified elsewhere; K73-Chronic hepatitis not classified elsewhere; K74-Liver fibrosis and cirrhosis; K75-Other inflammatory liver diseases; K76-Other liver diseases; K77-Liver disorders in diseases classified elsewhere.

During the 6-year investigation period, agricultural workers reported 382,869 YPLL, with 1,866,216 years reported among other workers. The proportional distribution of YPLL (% YPLL) by determinants showed higher proportions for both groups of workers in the K70 subgroup, among males, individuals of Brown race, and single individuals. Geographical analysis showed that 59.6% of YPLL among agricultural workers was significantly concentrated in the Northeast region, compared to only 22.6% among other workers in the same area (Table 4).

The national YPLLr for deaths associated with liver diseases was 3,119.9 years per 100,000 agricultural workers. Within this group, the Northeast regions notably had YPLLr of 4,527 years per 100,000 agricultural workers. Comparing the YPLLr ratio between agricultural and other workers, the rate for agricultural workers was 1.33 times higher nationally and 2.24 times higher in the Northeast region. Additionally, within the agricultural workers group, the Northeast region showed a significantly worse situation, with YPLLr ratio 2.78 times higher than the South region, which had the lowest YPLLr for liver diseases among agricultural workers (Table 5).

**Table 5.**
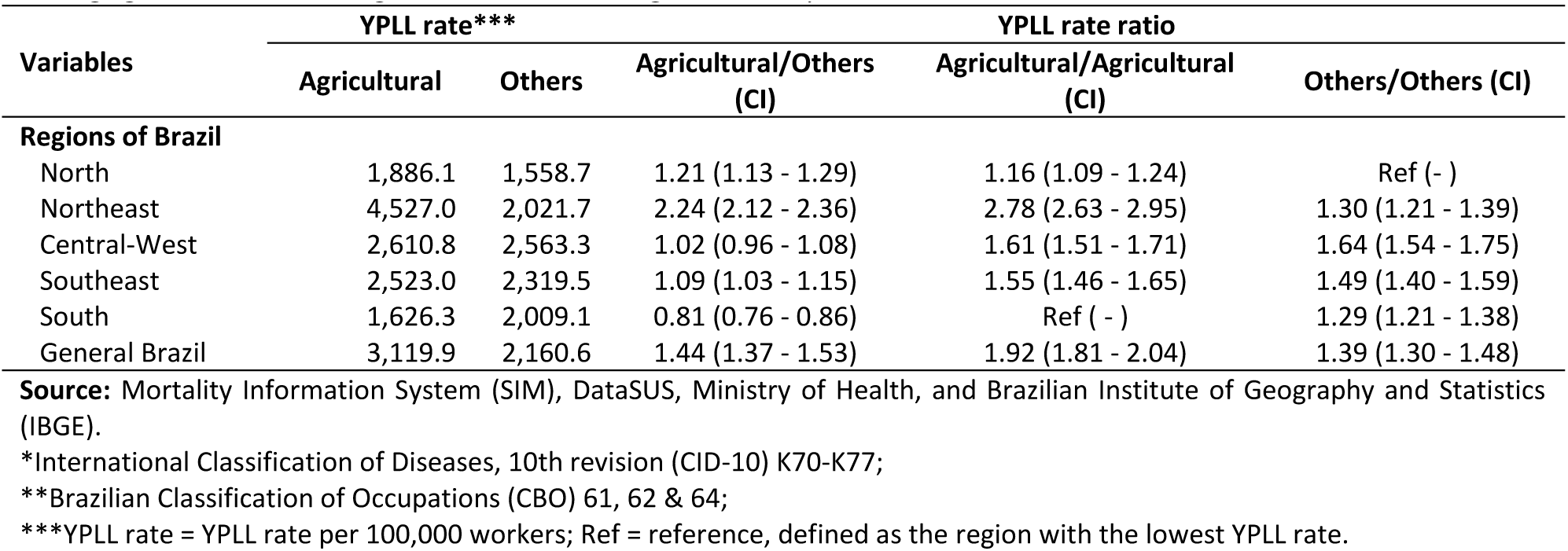
Rate and rate ratio with 95% confidence interval (CI) of Years of Potential Life Lost (YPLL) resulting from liver disease*, among agricultural and non-agricultural workers** aged 18 to 69 years in Brazil from 2017 to 2022.

## DISCUSSION

This is the first national study focusing on occupation and early deaths due to liver diseases among individuals aged 18 to 69. The early mortality, particularly among agricultural workers, is evident from the MAoD of 51.3 years and YPLLr of 4,532.5 per 100,000 agricultural workers during the study period. The type of liver disease identified and the concentration of deaths in Northeast Brazil, especially with higher proportions in the K70 subgroup - alcoholic liver disease, suggest explanatory hypotheses, such as possible early and excessive alcohol consumption, combined with a lack of access to preventive examinations or treatments during the early stages of liver diseases.

Diseases of the Digestive System (CID-10 - Chapter XI), was the 6^th^ leading underlying cause of death during the study period, accounting for approximately 227,000 deaths among Brazilians aged 18 to 69. Among these, liver diseases represented 52% of the deaths^34^. Pathologies of K70-K77 are challenging due to their multifactorial causes, including viral infections, poisoning by harmful substances, excessive alcohol consumption, high-fat diets, or genetic factors^3,6,7,12^. In the field of occupational health, efforts are focused on understanding the direct or indirect role of work in the development of these morbidities. For example, agricultural workers are highly exposed to exogenous agents that can cause or heighten chronic diseases^15^.

The population of agricultural workers in Brazil, the focus of this study, is approximately 15 million, predominantly male (81%), with 45% identified as White and 44% as Brown race, and low education level (70%). Economically, 70% are engaged in subsistence farming, primarily producing plant-based foods and raising animals for slaughter or their derivatives (milk, wool etc.)^35^. These workers, historically exploited and still facing significant social inequalities, endure routine long working hours, exposure to various pesticides and heavy metals, and vulnerability to weather conditions, among other hazards^15,23,24^. Such conditions increase their susceptibility to work-related accidents, diseases, and health issues ^7,36,37^. Additionally, considerable distances from standard healthcare centers pose challenges for agricultural workers in accessing preventive examinations or early treatment, leading to permanent disabilities or increased risk of death among them.

For comparison purposes, a reference group consisting of all other workers in the same age range and period was created. With no exclusion criteria applied, this group included all CBOs except agricultural workers, resulting in a heterogeneous mix regarding occupational risk or protective factors for liver diseases. Notably, the reference group had twice as many White workers, with higher levels of education, and a geographical distribution primarily concentrated in the Southeast and in urban municipalities. Epidemiologically, these differences are important for identifying potential determinants contributing to the outcome of interest.

The use of the YPLL indicator allows for qualifying and measuring the magnitude of deaths due to specific causes of interest. In occupational health^38–42^, YPLL permits quantification of the burden of work-related risks and, when feasible, presents determinants or exposure factors for diseases and injuries possibly associated with deaths. YPLL calculation is recommended when the data source has good quality dimensions, including coverage, completeness, reliability, and validity. SIM, the data source of this study, has satisfactory registration coverage for the occupation variable among Brazilian SIS (Sistemas de Informação da Saúde) despite some incompletely filled fields ^43^. In this study, the database had 83% completion rate for the occupation field, classified as regular in information quality^44^. Similarly, age, which is essential for YPLL calculation, has nearly 100% completion in the SIM records^45^.

In this study, the YPLL calculation used the LE of 76.2 years, based on the year 2019 for Brazil. This year was chosen as it represents the midpoint of the study period and the year before the onset of the COVID-19 pandemic, which eventually decreased LE in subsequent years. Here, the YPLL calculations did not consider differences in sex, race, or region where deaths occurred, as it is common in national and international studies to use a single LE value for YPLL calculations^39,41,42,46^. However, it is important to note that LE may vary by up to 10 years depending on regions or various other determinants. For instance, in 2019, LE for men and women was 73.1 and 80.1 years, and regionally, the North had the lowest LE at 72.9 years, while the South had the highest at 78.6 years. Similarly, the Northeast had LE of 73.9 years, the Midwest 75.8 years, and the Southeast 78.3 years ^33^. Historically, Black and Brown individuals usually have lower LE compared to White individuals^41,46–48^, likely due to centuries of inequality and systematic disadvantages. Therefore, the potential for under or overestimation of some results cannot be ruled out. Despite this methodological limitation, the use of data analysis techniques, such as rate and rate ratio calculations, are appropriate strategies for comparing different determinants and reducing potential biases.

For all calculated indicators, males exhibited significantly higher incidences, both among general and agricultural workers. These findings align with previous studies that consistently identify higher incidence and mortality rates from liver diseases in males, irrespective of occupation or other determinants^3,5,7,11^. This historical pattern of liver disease-related deaths may be attributed to several factors, including higher frequencies of risk behaviors such as excessive alcohol consumption and smoking, poor health-seeking behaviors, and greater environmental and occupational exposures^4,12,23^.

The 51.3 years MAoD resulting from liver diseases among agricultural workers highlights the early onset of this disease within the study population. In acute events such as work-related accidents, it is common for the average age at death to be low, often lower than these findings. For instance, among workers in the state of Bahia, the MAoD was 38.9 years for individuals of Brown race, 39.4 for Black individuals, and 40.6 for White individuals^41^. Similarly, a study among Iranian workers found MAoD of 36 years, which was higher than that among Turkish workers, who had MAoD of 34 years^40,49^. Mortality from non-communicable chronic diseases related to work, such as cancers and kidney diseases, typically have higher average age at death than liver disease-related deaths^50,51^. Premature deaths have profound impacts on families, causing disruption in the parental nucleus, loss of human references, and household income, resulting in economic loss for spouses and dependents. Socially, these deaths can exacerbate inequalities, reduce the country’s workforce, and negatively impact regional and national development^52^.

The low levels of education of agricultural workers in this study is similar to findings from previous national studies using primary data^37,53,54^ and the 2017 Agricultural Census^35^. A high proportion of agricultural workers have ≤ 7 years of schooling (52.4%), which is double that of other workers (27.3%), potentially contributing to the main findings here. When stratified by groups or regions, the low levels of education of agricultural workers, reveal specific vulnerabilities. For example, the percentage of agricultural workers who have never attended school is higher in states in the North and Northeast, which may be associated with the development of liver diseases^8,10^. This educational disparity may contribute to a lesser perception of behavioral or environmental risks, correlate with lower income, and result in poor health-seeking behaviors, leading to high morbidity and mortality from liver diseases.

Racially, the PM from liver diseases among Black agricultural workers (Brown race and Black individuals) was 19.4% higher compared to other Black workers (71.7% vs. 52.3%). Adverse occupational effects based on race are commonly investigated concerning workplace accidents. A national study published in 2021, using SIM data, identified that Brown race workers had the highest average annual mortality rates from workplace accidents across all regions of Brazil^55^. Additionally, a temporal trend analysis on fatal workplace accidents involving workers from Bahia between 2000 and 2019 concluded that Brown race workers had a lower MAoD (38.9 years), as well as higher average YPLL and higher YPLLr compared to Black and White race workers^41^. These findings are consistent with international literature, which identifies non-White and Hispanic workers as having a higher prevalence of injuries, assaults, and risks related to workplace accidents^56,57^.

Regarding the population of agricultural workers, an Epidemiological Bulletin from the Ministry of Health, using data from the Notifiable Diseases Information System, identified that 46.8% of accidents involving venomous animals and 45.4% of severe workplace accidents between 2010 and 2019 in Brazil occurred among Brown race agricultural workers^58^. Similarly, a systematic review with meta-analysis, evaluating 38 international studies (USA, Australia, Belgium, Canada, China, and Finland), identified higher risks of accidents and injuries among non-White agricultural workers^59^, which is similar to the principal findings of this study.

Despite the significant proportion of YPLL (73.8%) for liver diseases among Black and Brown individuals, YPLL rates were not calculated due to the limitation of lacking denominators related to occupation by race. As of May 2024, the main source of this data, IBGE did not provide tables with this information - Table 3584 (https://sidra.ibge.gov.br/Tabela/3584). Also, tables related to occupation with information by race did not allow tabulation by specific types of occupations, as seen with tables 4040 (https://sidra.ibge.gov.br/Tabela/4040) and 3581 (https://sidra.ibge.gov.br/Tabela/3581). Therefore, alongside determinants such as sex, age, and levels of education, location and occupation, there is an urgent need to include race/ethnicity in information systems, to permit widespread application of health indicators for workers.

The K70 subgroup - alcoholic liver disease was recorded as the underlying cause of death for 53.8% of the investigated agricultural workers. These findings align with a national retrospective historical series (1996-2022) that examined various liver-related morbidities, identifying alcoholic liver disease as the leading cause of death^12^. The K70 subgroup is primarily driven by chronic and high alcohol consumption^6,7,11^. This exposure can trigger the formation of stress granules and the restructuring of hepatic tissue, leading to the degeneration of various liver cells, including hepatocytes, stellate cells, cholangiocytes, and Kupffer cells^11^. The progressive inflammatory process results in the replacement of healthy hepatic tissue with nodules and fibrosis, compromising blood circulation^4^.

The relationship between alcohol consumption and agricultural workers has been extensively studied^15,20,23,24^. Potential determinants of abusive alcohol behavior include crop loss^24^, climate change^24^, low economic yields^23^, psychological adversities^23^, food insecurity^23^, intense and exhausting workload^23^, and high exposure to pesticides^15^. However, there is limited research on the consequences of this chronic exposure and its potential adverse health outcomes, such as the development of alcoholic liver disease, which was responsible for 8,276 deaths among Brazilian agricultural workers in the study period.

The unfavorable outcomes for liver disease-related deaths among agricultural workers in the Northeast region support previous studies that identified adverse conditions among workers in this region^36,39,41^. With approximately 68% of its municipalities classified as rural, the Northeast has the largest population of subsistence agricultural workers in the country^35^. Deficient infrastructure, such as basic health units, specialized centers, hospitals, and a lower number of healthcare professionals, especially in remote rural areas, exacerbates early morbidity and mortality among this population^36^. Consequently, there is a pressing need to expand occupational health surveillance and implement measures to improve working environments and conditions. Such actions are essential to reduce the burden associated with environmental and behavioral factors, like liver diseases and promote equity among the study population.

Reflecting on the limitations of the study, the YPLL calculation represents an absolute value, leading to higher YPLL values in large populations. To ensure comparability with studies and populations of different sizes, the YPLLr and YPLLr ratios were applied^41,60^, as these strategies are commonly used in epidemiological studies^42,46^. The YPLLr and YPLLr ratios confirmed the magnitude of the problem and highlighted the vulnerability of agricultural workers in the Northeast region of Brazil. Although no studies with directly comparable populations were found, these findings are similar to national studies that observed premature deaths among Brazilian workers due to chronic diseases^42^, infectious diseases^36^, or workplace accidents^38,41^.

Although this study relied on secondary data from SIM and IBGE, the quality of SIM records has improved significantly, particularly in the sociodemographic and occupational fields, which are now completed in 85% of the records. Additionally, there is a decreasing trend in the proportions of codes classified as ill-defined causes, especially within the investigated population aged 18 to 69 years.

The inability to measure variables such as alcohol consumption, use of non-prescription medications, dietary habits, and other factors limits the ability to infer causality of liver diseases among workers. Future epidemiological studies with prospective (cohort) and retrospective (case-control) designs, involving representative sample sizes of the worker population, are necessary to better assess the potential impact of multiple risk exposures, with an emphasis on alcohol consumption and its relation to early mortality from liver diseases among agricultural workers.

Highlighting the positive aspects of the study, approximately 4 million death records formed the initial basis of analysis. The application of indicators such as PM and YPLL, along with YPLLr and YPLLr ratio, enabled comparisons between groups of workers based on occupational, sociodemographic, and geographic aspects, paving the way for understanding liver disease mortality among agricultural workers in Brazil. The study’s findings prompt reflection on the determinants that have rendered this group of agricultural workers particularly vulnerable to liver diseases. The concentration of deaths in the K70 subgroup - Alcoholic liver disease suggests the need for developing continuous education strategies aimed at agricultural workers to raise awareness about the risks associated with early, high, and frequent alcohol consumption. Overall, this study contributes to the field of occupational health by fostering the development of targeted actions to promote the health and well-being of populations involved in agricultural activities.

## CONCLUSIONS

During the 6-year investigation period, 120,552 deaths associated with liver diseases were recorded among Brazilians aged 18 to 69 years. Of this, 12.7% occurred among agricultural workers, with MAoD of 51.3 years and a total of 382,869 YPLL. The higher proportion of deaths in the K70-Alcoholic liver disease subgroup suggests precocious, high, and frequent alcohol consumption among agricultural workers. The Northeast region had a YPLL rate of 4,527 years per 100,000 agricultural workers and YPLLr ratio 1.45 times higher than the national average, rendering it the region with the most severe situation nationwide. Characterizing the vulnerability of agricultural workers to liver diseases requires a combination of epidemiological methods to measure behavioral, environmental, and occupational risks. This approach should help fill the knowledge gaps not covered by the scope of this study.

## Data Availability

All datasets and codes used during this study are available in Zenodo under Creative Commons 4.0 license, accessible through https://doi.org/10.5281/zenodo.12093023

## Acknowledgements

J. S. Silva and T. S. Nunes received CNPq_IC Scientific Initiation and CAPES Master’s Scholarships during this study.

## Authors’ Contributions

J. S. Silva and C. Cremonese contributed equally to the conception of the study proposal; data collection; analysis; interpretation of results; drafting of the manuscript; and critical review of the final version. S. Arruda, T. S. Nunes, M. Armando and W. P. Dias performed data analysis; interpretation of results; and critical review of the manuscript. A. M. Awoniyi collaborated in the drafting of the manuscript; translation from Portuguese to English; and critical review of the manuscript. All authors read and approved the final version of the manuscript.

